# The MSPTDfast photoplethysmography beat detection algorithm: Design, benchmarking, and open-source distribution

**DOI:** 10.1101/2024.08.23.24312514

**Authors:** Peter H Charlton, Erick Javier Argüello-Prada, Jonathan Mant, Panicos A Kyriacou

## Abstract

**Objective:** Photoplethysmography is widely used for physiological monitoring, whether in clinical devices such as pulse oximeters, or consumer devices such as smart-watches. A key step in the analysis of photoplethysmogram (PPG) signals is detecting heartbeats. The MSPTD algorithm has been found to be one of the most accurate PPG beat detection algorithms, but is less computationally efficient than other algorithms. Therefore, the aim of this study was to develop a more efficient, open-source implementation of the MSPTD algorithm for PPG beat detection, named MSPTDfast (v.2).

**Approach:** Five potential improvements to MSPTD were identified and evaluated on four datasets. MSPTDfast (v.2) was designed by incorporating each improvement which on its own reduced execution time whilst maintaining a high F_1_-score. After internal validation, MSPTDfast (v.2) was benchmarked against state-of-the-art beat detection algorithms on four additional datasets.

**Main results:** MSPTDfast (v.2) incorporated two key improvements: pre-processing PPG signals to reduce the sampling frequency to 20 Hz; and only calculating scalogram scales corresponding to heart rates >30 bpm. During internal validation MSPTDfast (v.2) was found to have an execution time of between approximately one-third and one-twentieth of MSPTD, and a comparable F_1_-score. During benchmarking MSPTDfast (v.2) was found to have the highest F_1_-score alongside MSPTD, and amongst one of the lowest execution times with only MSPTDfast (v.1), qppgfast and MMPD (v.2) achieving shorter execution times.

**Significance:** MSPTDfast (v.2) is an accurate and efficient PPG beat detection algorithm, available in an open-source Matlab toolbox.

## 1. Introduction

Photoplethysmography sensors are widely used in physiological measurement, whether in clinical devices such as pulse oximeters, or consumer devices such as smart-watches [1]. Photoplethysmography is a non-invasive optical sensing technology, which measures the arterial pulse wave [2]. A wide range of applications of photoplethysmography have been proposed, from cardiovascular to respiratory to mental health monitoring [3]. Each application relies on accurate extraction of physiological information from the photoplethysmogram (PPG) signal. A key step in extracting physiological information from the PPG is the detection of individual pulse waves, corresponding to heartbeats [4]. Consequently, it is important to develop accurate and efficient algorithms for PPG beat detection.

A wide range of PPG beat detection algorithms have been proposed [5], and open-source implementations are available for several of these algorithms [6]. A recent benchmarking study assessed the performance of open-source PPG beat detection algorithms, and identified the *MSPTD* and *qppgfast* algorithms as the best-performing algorithms [6]. However, this study only assessed the accuracy of the algorithms, and did not assess their efficiency. Here, efficiency can be thought of in terms of the time required to run an algorithm, and its memory requirements [7]. Efficiency is an important aspect of algorithm performance alongside beat detection accuracy, because it directly impacts the range of devices an algorithm can be deployed on, as well as the device’s power consumption. Pulse oximeters and wearable devices operate under resource constraints, and so only sufficiently efficient algorithms can be used on these devices. Furthermore, an algorithm’s efficiency affects a device’s power consumption, which in turn determines influences how frequently PPG measurements can be taken whilst maintaining the desired battery life. In initial experiments we observed that the *MSPTD* algorithm had a much longer execution time than other leading algorithms (such as *qppgfast*), indicating that it could be beneficial to increase its efficiency [8].

The aim of this work was to develop a more efficient, open-source implementation of the *MSPTD* algorithm for PPG beat detection, named *MSPTDfast (v*.*2)*. The objectives were to: (i) design *MSPTDfast (v*.*2)* by evaluating its accuracy and performance when including each of several potential improvements; (ii) benchmark *MSPTDfast (v*.*2)* against leading beat detection algorithms; and (iii) distribute and validate *MSPTDfast (v*.*2)* in an open-source toolbox. This work builds on a recent conference paper [8], which presented an initial design for *MSPTDfast (v*.*1)* developed using a subset of one of the eight datasets used in the present study, and benchmarked against two of the six comparator algorithms used in the present study.

## 2. Methods

The experiments reported in this study used the *ppg-beats* framework [9] developed in [6] to assess the performance of PPG beat detection algorithms. This framework includes benchmark datasets, implementations of state-of-the-art algorithms, and evaluation code. The framework has been improved and extended since its original description, with key changes highlighted in this section. Some aspects of the Methods and Section 3.1.1 are reproduced from [8] under [CC BY 4.0](https://creativecommons.org/licenses/by/4.0/).

Ethical approval was not required for this study as it used pre-existing, anonymised data.

### 2.1. Datasets

A total of eight freely available datasets were used in this study, as summarised in Table 1. Each dataset includes PPG signals and simultaneous ECG signals from which to obtain timings of reference heartbeats. The datasets cover a range of scenarios, from finger PPG recordings in hospital patients to wrist PPG recordings in healthy volunteers.

**Table 1.** Datasets used to assess the performance of PPG beat detectors. *Source: adapted from [6] under CC BY 4.0*.

| Dataset | Subjects | PPG equipment | Reference beats | Duration (mins): med (quartiles) | Total beats |
| --- | --- | --- | --- | --- | --- |
| *Development datasets* |  |  |  |  |  |
| CapnoBase | 42 patients undergoing elective surgery and routine anaesthesia [10]. | Pulse oximeter at 300 Hz (upsampled from 100 Hz during acquisition) | Manual annotations of ECG (300 Hz) | 7.7 (7.0 - 7.7) | 24,953 |
| BIDMC | 53 critically-ill adult patients, a subset of the MIMIC II dataset [11]. | Bedside monitor at 125 Hz (mostly finger PPG recordings) | ECG-derived QRS detections (125 Hz) | 7.4 (7.0 - 7.7) | 33,520 |
| MIMIC PER-form Training Dataset | 200 critically-ill patients during routine clinical care (100 adults, 100 neonates) [6]. | Bedside monitor at 125 Hz (mostly finger PPG recordings) | ECG-derived QRS detections (125 Hz) | 6.2 (4.1 - 8.2) | 129,708 |
| PPG-DaLiA | 15 subjects during a protocol of activities of daily living [12]. | Wristband (Empatica E4) at 64 Hz (with corresponding tri-axial accelerometry signals). | Manual annotations of ECG (700 Hz) | 139.4 (129.8 - 147.3) | 183,941 |
| *Testing datasets* |  |  |  |  |  |
| WESAD | 15 subjects during a laboratory-based protocol designed to induce different emotions [13]. | Wristband (Empatica E4) at 64 Hz (with corresponding tri-axial accelerometry signals). | ECG-derived QRS detections (700 Hz) | 87.8 (83.2 - 92.4) | 103,222 |
| MIMIC PER-form Testing Dataset | 200 critically-ill patients during routine clinical care (100 adults, 100 neonates) [6]. | Bedside monitor at 125 Hz (mostly finger PPG recordings) | ECG-derived QRS detections (125 Hz) | All: 6.2 (3.8 - 8.4)<br>Adults: 8.3 (6.5 - 9.3)<br>Neonates: 4.3 (3.0 - 5.8) | All: 127,383<br>Adults: 64,280<br>Neonates: 63,103 |
| MIMIC PER-form AF Dataset | 35 critically-ill adults during routine clinical care (19 in AF, 16 not in AF), using AF labels provided by cardiologists [14, 15]. | Bedside monitor at 125 Hz (mostly finger PPG recordings) | ECG-derived QRS detections (125 Hz) | AF: 19.1 (17.3 - 19.7)<br>non-AF: 19.1 (18.7 - 19.7) | AF: 31,929<br>non-AF: 23,572 |
| MIMIC PER-form Ethnicity Dataset | 200 critically-ill adults during routine clinical care (100 of Black ethnicity, 100 of White) [6]. | Bedside monitor at 125 Hz (mostly finger PPG recordings) | ECG-derived QRS detections (125 Hz) | Black: 8.6 (6.8 - 9.4)<br>White: 8.2 (5.9 - 9.1) | Black: 68,214<br>White: 62,054 |

#### 2.1.1. Development datasets

Four datasets were used in the development of *MSPTDfast (v*.*2)*: CapnoBase, BIDMC, MIMIC PERform (Training), and PPG-DaLiA. The CapnoBase, BIDMC and MIMIC PERform (Training) datasets contain predominantly high-quality PPG signals acquired via pulse oximeters in hospital. In contrast, the PPG-DaLiA dataset contains lower-quality PPG signals acquired from wearables in activities of daily living. The CapnoBase dataset contains recordings from 29 paediatric and 13 adult patients during surgery and anaesthesia [10]. The BIDMC dataset contains recordings from 53 critically-ill adult patients [11]. The MIMIC PERform (Training) dataset contains recordings from 200 critically-ill patients (100 adults, and 100 neonates) [6]. BIDMC and MIMIC PERform (Training) were both extracted from the larger MIMIC database [6]. The PPG-DaLiA dataset contains recordings acquired using the wrist-worn Empatica E4 device from 15 subjects during a protocol of activities of daily living (the activities consisted of sitting, working, cycling, walking, on a lunch break, car driving, stair climbing, and playing table soccer) [12]. The subjects were mostly young adults, with a median (lower-upper quartiles) age of 28 (24–36) years.

#### 2.1.2. Testing datasets

Four datasets were used to test *MSPTDfast (v*.*2)* and other beat detection algorithms. Of these, the MIMIC PERform (Testing) and WESAD datasets were used to test performance. The MIMIC PERform (Testing) dataset is similar to the previously described MIMIC PERform (Training) dataset: it contains PPG signals acquired via pulse oximeters from a different set of 200 critically-ill patients (100 adults and 100 neonates) [6]. The WESAD dataset is similar to PPG-DaLiA in that it also contains recordings acquired using the wrist-worn Empatica E4 device from subjects during a research protocol, in this case a protocol designed to induce different emotions (consisting of a baseline measurement, and measurements during meditation, amusement, and stress) [13]. In addition, the MIMIC PERform Testing, AF and Ethnicity datasets were used to investigate associations between the performance of beat detection algorithms and patient characteristics: between 100 adults and 100 neonates (Testing); between 19 subjects with and 16 subjects without atrial fibrillation (AF); and between 100 Black and 100 White subjects (Ethnicity).

It should be noted that there were two key changes to the way in which the datasets were used since the *ppg-beats* assessment framework was originally described in [6]. First, the PPG-DaLiA dataset was used for algorithm development and the WESAD dataset for testing, whereas the opposite was recommended in the original description. This was because part of the PPG-DaLiA dataset had been used for initial development of *MSPTDfast (v*.*1)* in [8], and thus avoided any overfitting. Second, the entire PPG-DaLiA and WESAD datasets were used in this analysis rather than separating these datasets into subsets according to activities. This approach was taken to simplify the presentation and interpretation of results. It means that not only periods of specific activities were included in the analysis (as was the case previously), but in addition periods between protocolised activities were also included.

### 2.2. Designing *MSPTDfast (v*.*2)*

#### 2.2.1. The *MSPTD* Algorithm

The original Multi-Scale Peak & Trough Detection (*MSPTD*) algorithm [16] was a refinement of the Automatic Multiscale Peak Detection (*AMPD*) algorithm [17]. Both of these algorithms identify peaks in a signal as points which are higher than their surrounding neighbours, where the number of surrounding neighbours to consider is determined through analysis of a local maxima scalogram. The local maxima scalogram is a 2D matrix where each element indicates whether the current sample is higher than its two neighbours at a certain scale. For instance, the top row indicates whether points are higher than both their immediate neighbours (a separation of 1 sample), the second row indicates whether points are higher than their next nearest neighbours (a separation of 2 samples), etc. For a point to be classified as a peak, it must be higher than its neighbours on all rows of the scalogram until a certain scale (*i*.*e*. level of separation). This scale is identified as the row in which most points are classified as peaks. Further details of the algorithms are provided in Figure 1. The *MSPTD* algorithm was developed to improve the computational efficiency of *AMPD*. This improvement was achieved by using binary values rather than random numbers in the scalogram. It was also modified to detect both peaks and onsets. For further explanation see [18].

**Figure 1.**
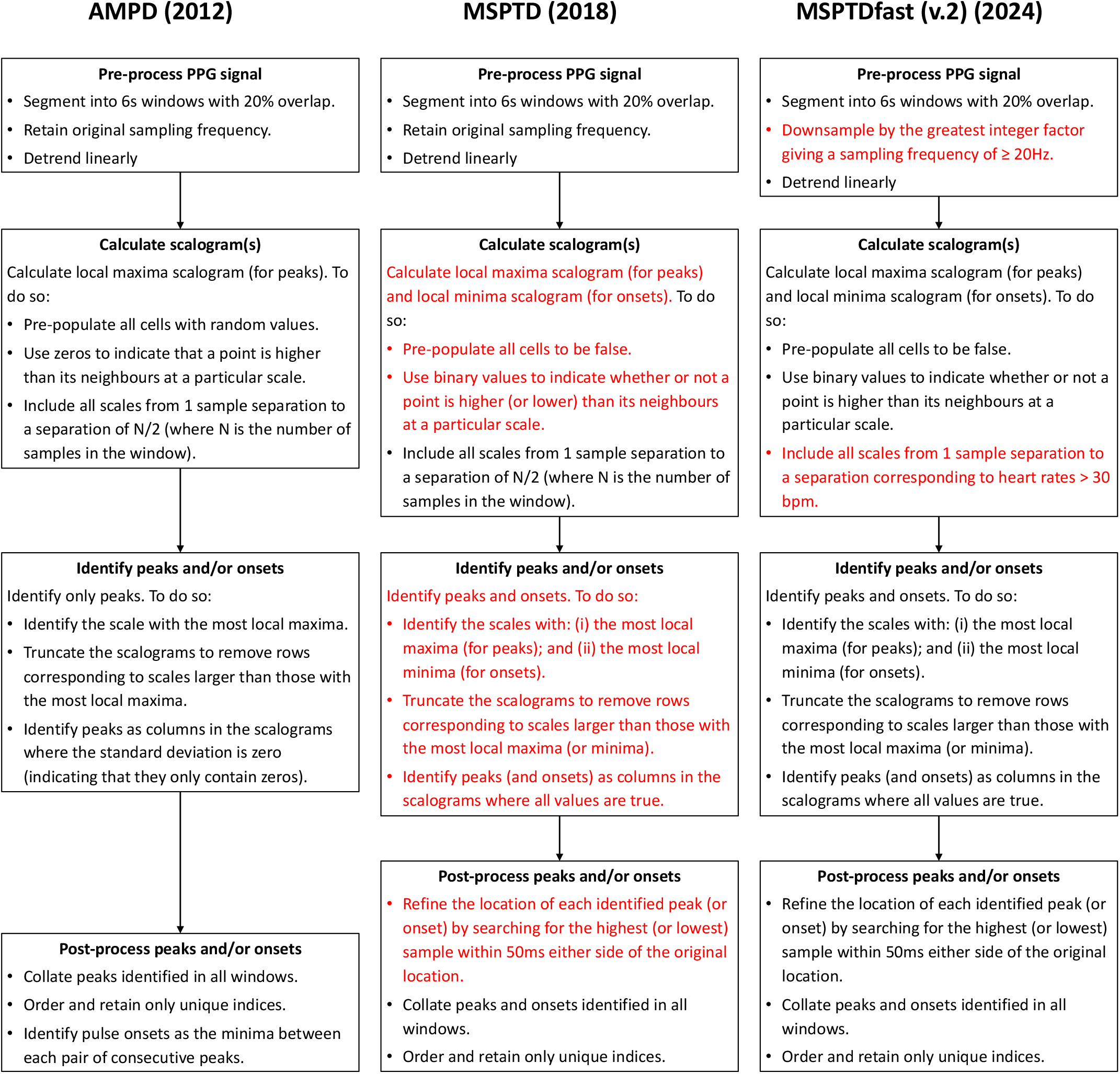
The development of *MSPTDfast (v.2)*. The flowcharts show the main steps in three PPG beat detection algorithms (as implemented in the *ppg-beats* toolbox [9]), with changes from one algorithm to the next shown in red. The algorithms are: (i) the Automatic Multiscale Peak Detection (*AMPD*) algorithm [17]; (ii) the Multi-Scale Peak & Trough Detection (*MSPTD*) algorithm [16], which improved the efficiency of *AMPD*; and (iii) the *MSPTDfast (v.2)* algorithm [this publication], which improved the efficiency of *MSPTD*.

#### 2.2.2. Improving *MSPTD*

We identified several potential ways to increase the efficiency of *MSPTD* through critically reviewing its current implementation (as described in [8]):

- **Number of scalograms calculated:** The *MSPTD* algorithm calculates scalograms corresponding to both pulse wave peaks and onsets. It may be possible to maintain beat detection accuracy whilst only calculating one scolgram, corresponding to either pulse wave peaks or onsets.
- **Method for calculating scalograms:** It has previously been proposed that the scalograms could be calculated more efficiently by vectorising the calculation method, thus avoiding the computationally expensive nested for loops used in *MSPTD* [18].
- **Number of scales used in each scalogram:** The scalograms calculated in *MSPTD* include all scales corresponding to levels of separation between the point of interest and its neighbours from one sample to *N/*2 samples (where *N* is the length of the signal). It may be possible to maintain beat detection accuracy whilst using a reduced set of scales, excluding those corresponding to separations longer than that between consecutive beats at a minimum plausible heart rate, *HR*_*min*_.
- **PPG sampling frequency:** PPG signals are typically sampled at >50 Hz, even though the signals are often filtered to exclude frequency content above 8-25 Hz prior to analysis [19]. Therefore, it may be possible to downsample the PPG signal prior to beat detection whilst still retaining the pertinent information on individual heartbeats (given that heart rates are rarely above 200 bpm [20, 21]).
- **PPG window duration:** Adjust the PPG window duration from the value of 6 s used in *MSPTD*.

Each of these potential improvements is aimed at increasing the efficiency of calculating the scalograms, since this is the most computationally complex part of the *MSPTD* algorithm.

The configuration options trialled for each potential improvement are summarised in Table 2. We assessed the impact of each option on the performance of a refined *MSPTD* algorithm in turn, whilst all other potential improvements were set to their default configuration options. The *MSPTDfast (v.2)* algorithm was designed by selecting each configuration option which provided the shortest execution time whilst maintaining a reasonably high *F*_1_-score (a subjective process). Its performance was internally validated through comparison with the original *MSPTD* algorithm on the development datasets.

**Table 2.** Evaluated algorithm configurations, where * indicates a default option. *Source: adapted from [8] under CC BY 4.0*.

| Potential improvement | Configuration options |
| --- | --- |
| Number of scalograms calculated | peaks; onsets; peaks and onsets * |
| Method for calculating scalograms | nested for loops *; vectorised approach |
| Number of scales used in each scalogram | $N/2$ *; only scales $> HR_{min}$, with $HR_{min} \in \{30, 40\}$ bpm. |
| PPG sampling frequency (Hz) | 10; 20; 30; original * |
| PPG window duration (s) | 4; 6; 8 *; 10 |

### 2.3. Benchmarking against leading beat detection algorithms

*MSPTDfast (v.2)* was implemented and distributed in the open-source *ppg-beats* toolbox [9], and benchmarked against state-of-the-art beat detection algorithms. The comparator state-of-the-art beat detection algorithms are summarised in Table 3. These consisted of: (i) the four best-performing algorithms identified in a previous benchmarking assessment [6] (*MSPTD* [16], *qppgfast* [22], *ABD* [23], and *AMPD* [17]); (ii) *MSPTDfast (v.1)* [8]; (iii) a new implementation of the ‘Mountaineer’s Method for Peak Detection’ algorithm (*MMPD (v.2)*) provided by the original author (whereas the version assessed in [6] was implemented by someone else) ^1^; and (iv) the recently reported ‘Waveform Envelope Peak Detection’ (*WEPD*) beat detector [24].

**Table 3.**
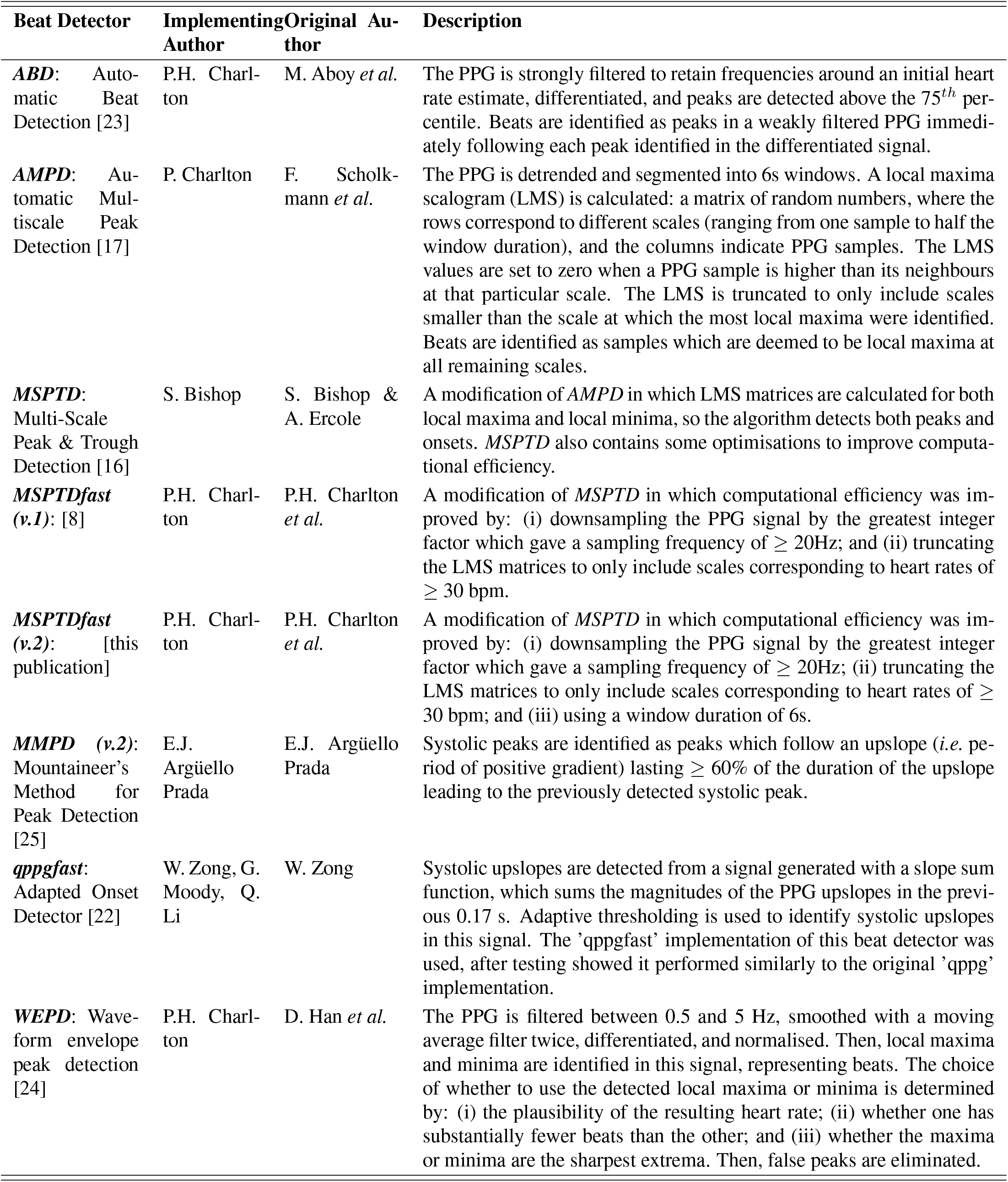
PPG Beat Detection Algorithms. *Source: adapted from [6] under CC BY 4.0*.

### 2.4. Performance Analysis

Performance was assessed in terms of sensitivity, positive predictive value, and the *F*_1_-score, indicating the algorithm’s ability to detect beats correctly. The computational efficiency was assessed using the algorithm execution time, indicating its efficiency. The methods used to calculate these metrics are now described.

Beats were detected in PPG signals using each PPG beat detection algorithm as follows. First, PPG signals were segmented into 20 s windows with 5 s overlap. Second, PPG signals were band-pass filtered between 0.67 and 8.00 Hz. Third, beats were detected using a given PPG beat detection algorithm. Fourth, the detected beats were post-processed by: (i) removing any repeated beat detections (arising in part due to the overlapping windows); and (ii) tidying up detected peaks and onsets to make them logically consistent with each other (ensuring that no peaks and onsets occurred at the same time; there was at least one local minimum between consecutive peaks, and at least one local maximum between consecutive onsets; that detections alternated between onsets and peaks; that detections started with an onset, and ended with a peak; and that there was the same number of peaks and onsets). Fifth, mid-points on PPG pulse wave systolic upslopes were detected between onsets and subsequent peaks, and used for analysis (because this point has been found to be optimal for pulse rate variability analysis [26]). Finally, any periods of no PPG signal were excluded from the analysis (*e.g*. due to missing values or flat line segments).

Reference beats were detected in ECG signals as follows. First, ECG signals were pre-processed by segmenting into windows in the same manner as for beat detection in PPG signals, and high-pass filtered above 0.67 Hz to remove baseline wander. Second, beats were detected in ECG signals using two ECG beat detectors: the *jqrs* [27, 28] and *RDECO* [29, 30] beat detectors. Only beat detections were identified as those which were agreed on by both beat detectors (*i.e*. within 150ms of each other) were retained for analysis.

Beats in PPG and ECG signals were time-aligned as follows to account for PPG and ECG clocks not being perfectly synchronised. The time-lag between the signals was identified as that which resulted in the highest number of agreed ECG and PPG beat detections (*i.e*. within 150ms of each other), with time-lags of between -10 and 10s trialled (with increments of 0.02s). In a refinement to the *ppg-beats* assessment framework, inspired by [31], the time-lag was allowed to vary within a recording to account for drifting clocks (which was found to be particularly important on the WESAD dataset), with new time-lags calculated every 300s.

Performance metrics were calculated for each subject’s recording as described in [6]. Reference beats were determined to be correctly identified if the closest PPG-derived beat was within ±150 ms of a reference beat. Then, the numbers of reference beats (*n*_ref_), PPG-derived beats (*n*_PPG_), and correctly identified beats (*n*_correct_) were used to calculate the following:

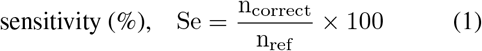

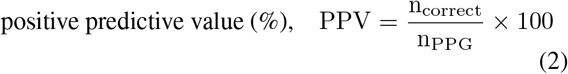

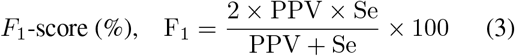

Execution time was calculated as the time taken to run the algorithm on a computer (in this case a MacBook Air M1 2020 without parallelisation), expressed as a percentage of the PPG signal duration. Performance metrics were expressed using the median and interquartile range, with boxplot whiskers showing 10^th^ and 90^th^ percentiles. The performance of *MSPTDfast (v.2)* was compared to that of other leading beat detection algorithms using the Wilcoxon rank sum test. Correction for multiple comparisons was performed using a Holm-Sidak correction [32, 33], for *F*_1_-scores of associations between beat detection accuracy and patient characteristics.

All analyses were performed in MATLAB (The Mathworks, Natick, MA, USA).

## 3. Results

### 3.1. Design

#### 3.1.1. Evaluating and selecting potential improvements

Figure 2 presents the performance of *MSPTDfast (v.2)* when using each of the potential improvements independently. Results are shown in terms of beat detection accuracy (median *F*_1_-score in red), and efficiency (median execution time in blue). Results are presented for each potential improvement in rows (a)-(e), with the plots in each row showing the results on each of the development datasets. Each potential improvement is now discussed in turn.

**Figure 2.**
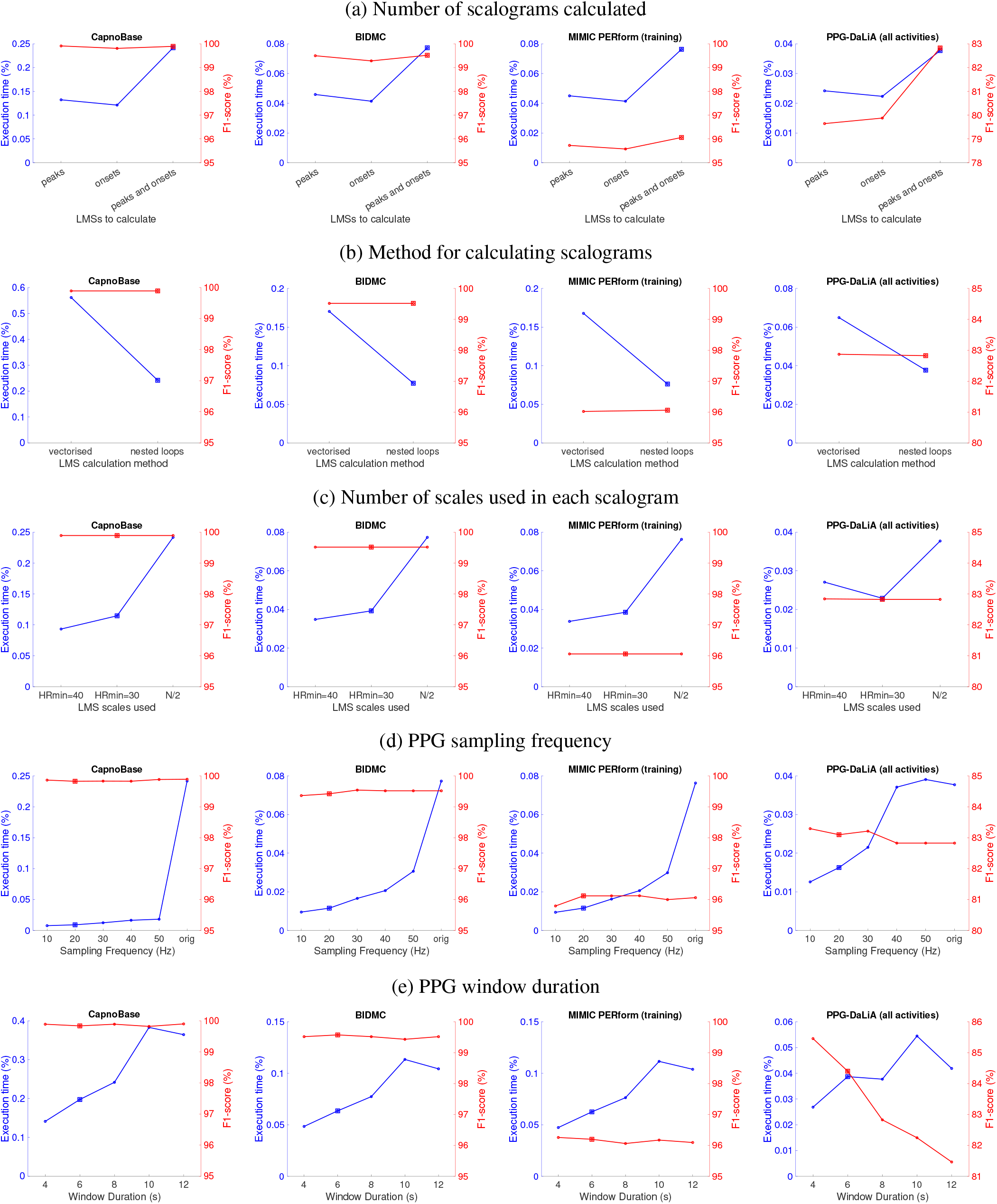
The performance of different algorithm configurations: (a)-(e) show performance when using different potential improvements, where squares indicate the configurations used in *MSPTDfast (v.2)*.

Calculating only a single LMS corresponding to either peaks or onsets consistently resulted in substantial reductions in execution time of between 36% and 50%, compared to calculating LMSs for both peaks and onsets (see (a)). However, this was accompanied by substantial reductions in *F*_1_-score on the PPG-DaLiA dataset of 3.2% when only calculating a LMS for peaks, and 2.9% when only calculating a LMS for onsets. Therefore, the ‘peaks and onsets’ option was selected for *MSPTDfast (v.2)* as the reduction in execution time achieved when only detecting peaks or onsets was not considered worthwhile when accompanied by a reduction in *F*_1_-score.

When comparing methods for calculating the LMSs, there was a substantial increase in execution time across all datasets when using our implementation of a vectorised approach in comparison to the original ‘nested loops’ approach (see (b)). Therefore the original ‘nested loops’ approach was retained for *MSPTDfast (v.2)*.

Reducing the number of LMS scales by eliminating scales corresponding to HRs below 30 or 40 bpm substantially reduced the execution time across all datasets (see (c)). Eliminating scales corresponding to HRs below 30 (or 40) bpm reduced execution time by 39% to 53% (or 28% to 61%). Whilst in this analysis there was no reduction in *F*_1_-score when using either minimum HR value, we selected the more conservative *HR*_*min*_ = 30 bpm for *MSPTDfast (v.2)* to retain accuracy at low heart rates.

Reducing the sampling frequency substantially reduced the execution time across all datasets (see (d)). Reducing the sampling frequency to 10Hz reduced execution time by between 67 and 97%. This was accompanied by minimal changes in *F*_1_-score, or even a slight increase in the case of the PPG-DaLiA dataset. We selected to downsample signals to 20 Hz in *MSPTDfast (v.2)*. We selected 20 Hz rather than the even more efficient 10 Hz, in an attempt to avoid reduced accuracy at high heart rates in neonates.

Reducing the window duration generally reduced execution time, whilst increasing the *F*_1_-score (see (e)). A duration of 6 s was selected for *MSPTDfast (v.2)*. This was selected rather than the even more favourable 4 s duration in an attempt to avoid reduced accuracy at low heart rates. In summary, the chosen configuration for *MSPTDfast (v.2)* was quite different to that of *MSPTD*: the number of LMS scales calculated was reduced by only calculating those scales corresponding to HRs of over 30 bpm; and the width of the LMS matrices was reduced by reducing the sampling frequency. *MSPTDfast (v.2)* also differed slightly to that of *MSPTDfast (v.1)*, in that a window duration of 6 s was used instead of 8 s. Figure 1 shows the main changes between *MSPTD* and *MSPTDfast (v.2)*.

#### 3.1.2. Internal validation

Figure 3 shows a comparison of *MSPTDfast (v.2)* and *MSPTD* on the development datasets in terms of the beat detection accuracy and efficiency. Corresponding results for sensitivity and positive predictive value are shown in Appendix A. The results demonstrate that the chosen configuration for *MSPTDfast (v.2)* resulted in the desired improvements over *MSPTD*: increased efficiency whilst maintaining accuracy. *MSPTDfast (v.2)* had an execution time of between approximately one-third and one-twentieth of the *MSPTD* algorithm: *MSPTDfast (v.2)*’s execution time was 5.3% of that of *MSPTD* on CapnoBase; 15.1% on BIDMC; 14.3% on MIMIC PERform (Training); and 35.9% on PPG-DaLiA. This was accompanied by minimal differences in *F*_1_-score: no change on CapnoBase; a decrease of 0.1% on BIDMC; an decrease of 0.2% on MIMIC PERform (Training); and an increase of 0.2% on PPG-DaLiA.

**Figure 3.**
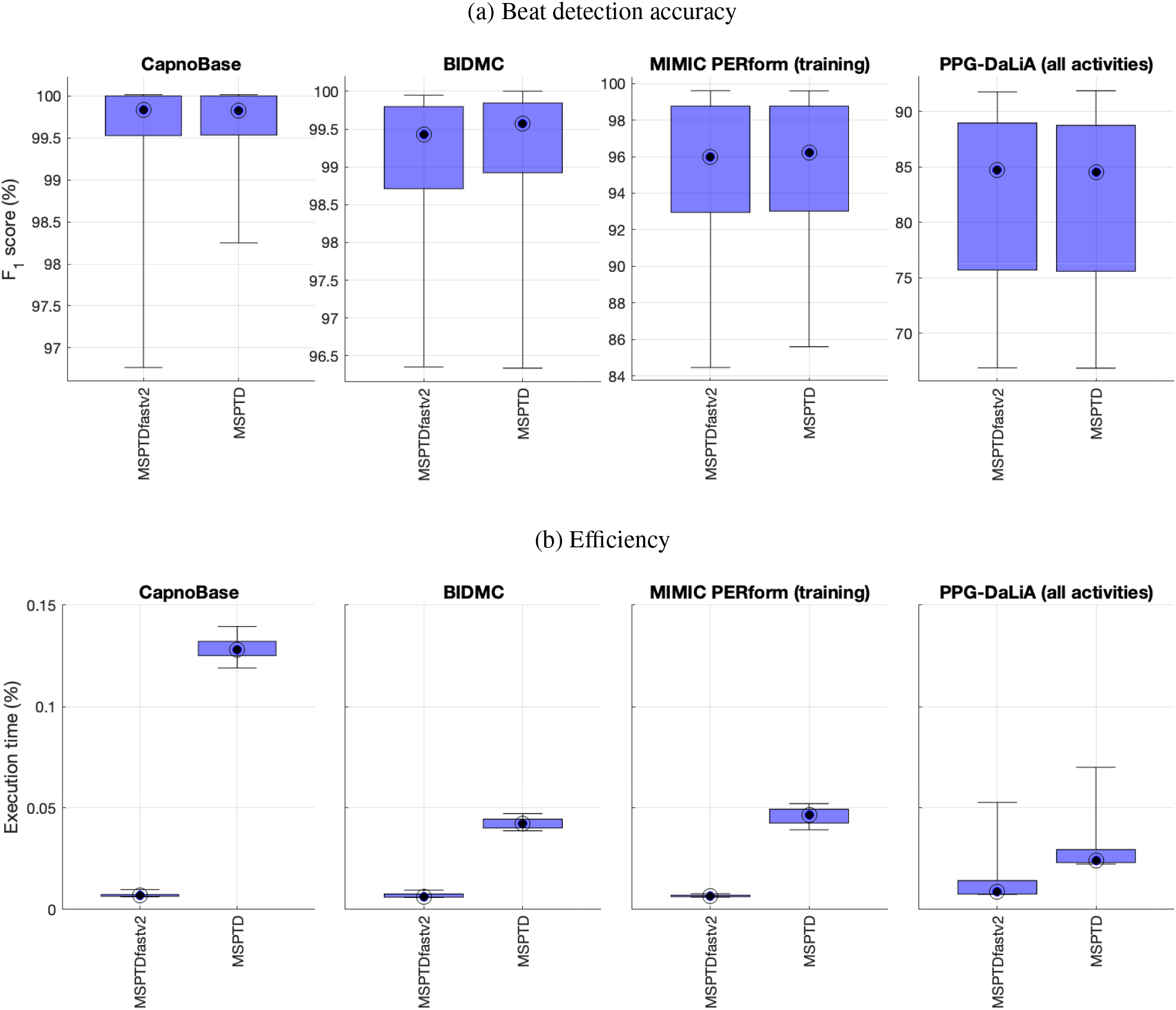
Internal validation of *MSPTDfast (v.2)* against *MSPTD* on the development datasets: Performance is shown in terms of (a) beat detection accuracy; and (b) efficiency. Corresponding results for sensitivity and positive predictive value are shown in Appendix A.

### 3.2. Benchmarking

The performance of the *MSPTDfast (v.2)* algorithm is compared to that of leading beat detection algorithms on the test datasets in Figure 4 and Table 4. Corresponding results for sensitivity and positive predictive value are provided in Appendix B. *MSPTDfast (v.2)* and *MSPTD* achieved the highest *F*_1_-scores on both datasets, and in both cases there was no significant difference between their *F*_1_-scores. *MSPTDfast (v.2)* achieved *F*_1_-scores of 96.8 (91.1 - 98.8) % and 84.6 (80.3-87.4) % on MIMIC PERform (Testing) and WESAD respectively. *MSPTDfast (v.2)* was also amongst the most efficient algorithms, with only *MSPTDfast (v.1)* achieving a shorter execution time on MIMIC PERform (Testing), and *qppgfast* and *MMPD (v.2)* achieving shorter execution times on WESAD.

**Table 4.** Benchmarking *MSPTDfast (v.2)* against leading beat detection algorithms. Results are expressed as median (lower - upper quartiles). Corresponding results for sensitivity and positive predictive value are shown in Appendix B. *Definition: * indicates a significant difference compared to MSPTDfast (v.2)*.

| Algorithm | MIMIC PERform (Testing) |  | WESAD (all activities) |  |
| --- | --- | --- | --- | --- |
| | $F_1$-score (%) | Execution time (%) | $F_1$-score (%) | Execution time (%) |
| MSPTD | 96.8 (90.8-98.9) | 0.0491 (0.0444-0.0558) * | 84.6 (80.3-87.5) | 0.0256 (0.0240-0.0319) * |
| MSPTDfastv2 | 96.8 (91.1-98.8) | 0.0074 (0.0066-0.0091) | 84.6 (80.3-87.4) | 0.0074 (0.0065-0.0136) |
| AMPD | 96.6 (90.2-98.8) * | 0.0587 (0.0537-0.0635) * | 83.3 (78.9-85.9) * | 0.0221 (0.0189-0.0260) * |
| MSPTDfastv1 | 96.6 (90.6-98.6) * | 0.0071 (0.0064-0.0086) * | 84.3 (80.2-87.2) * | 0.0071 (0.0066-0.0134) |
| qppgfast | 95.9 (89.5-98.6) * | 0.0078 (0.0070-0.0091) * | 82.9 (75.7-86.4) * | 0.0058 (0.0051-0.0123) * |
| ABD | 95.5 (88.6-98.1) * | 0.4396 (0.4066-0.4998) * | 83.2 (78.7-85.6) * | 0.4168 (0.3992-0.4333) * |
| MMPDv2 | 93.3 (87.6-97.6) * | 0.0092 (0.0084-0.0107) * | 84.1 (80.8-87.1) | 0.0072 (0.0064-0.0133) * |
| WEPD | 90.1 (75.1-97.5) * | 0.0250 (0.0227-0.0274) * | 83.5 (78.9-86.3) * | 0.0161 (0.0151-0.0215) * |

**Figure 4.**
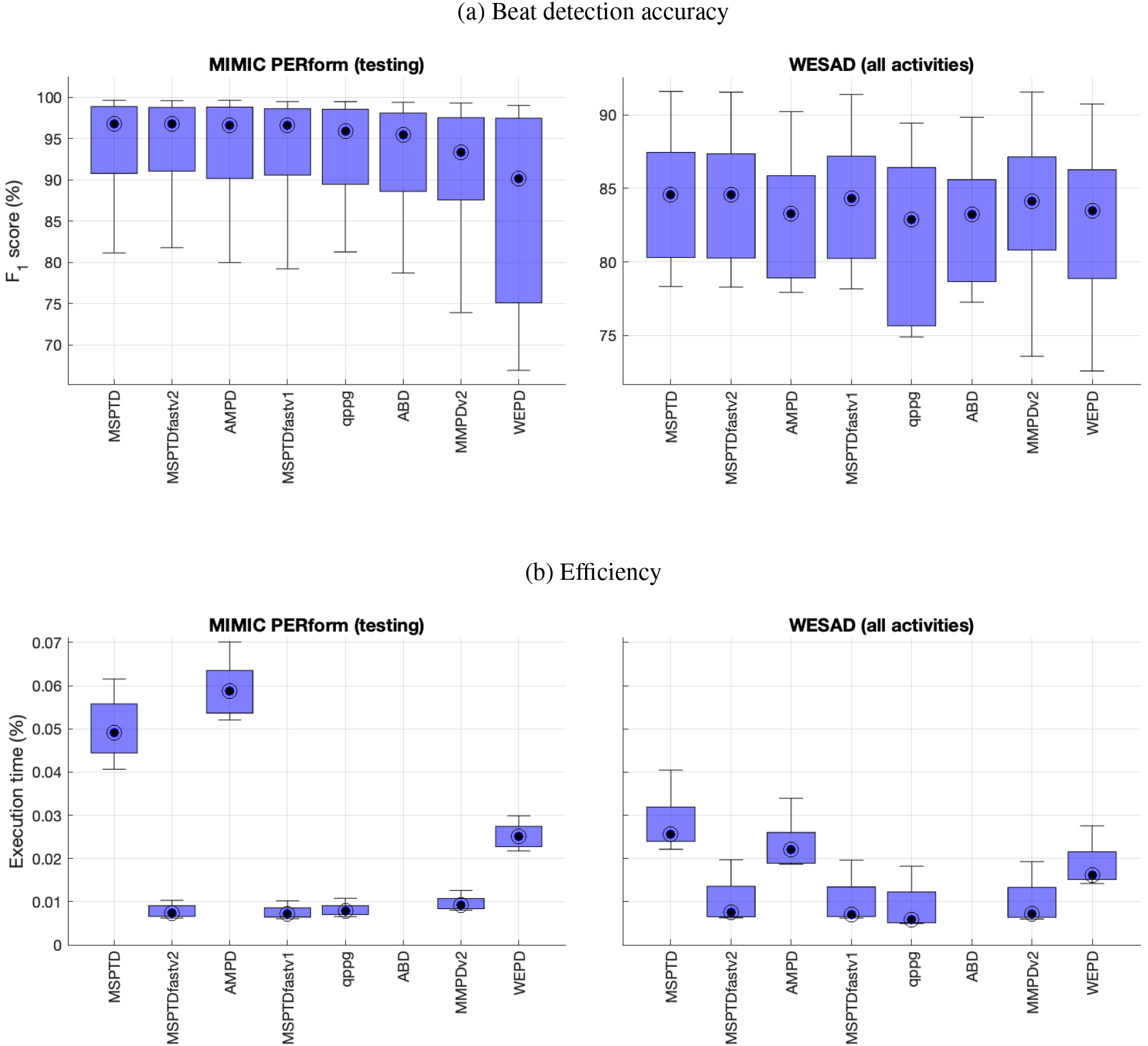
Benchmarking *MSPTDfast (v.2)* against leading beat detection algorithms: Performance is shown in terms of (a) beat detection accuracy; and (b) efficiency. Corresponding results for sensitivity and positive predictive value are shown in Appendix B.

The results also provide insight into the performance of other state-of-the-art algorithms. Generally, those algorithms which are based on the original *AMPD* method (*i.e. AMPD, MSPTD, MSPTDfast (v.1)*, and *MSPTDfast (v.2)*) achieved the highest *F*_1_-scores on both datasets, with the one exception that *MMPD (v.2)* also performed well on WESAD. An algorithm could achieve a relatively high *F*_1_-score on one dataset but a relatively low *F*_1_-score on the other dataset. For instance, *MMPD (v.2)*’s achieved a relatively high *F*_1_-score on WESAD (which was not significantly different to the highest *F*_1_-scores), but achieved the second lowest *F*_1_-score on MIMIC PERform (Testing). *WEPD* also showed quite different performance across the two datasets. In contrast, several different approaches achieved low execution times: the *MSPTDfast* algorithms, *qppgfast*, and *MMPD (v.2)* all had substantially lower execution times on both datasets than the remaining algorithms.

Figure 5 shows the associations between beat detection accuracy and patient characteristics. These results highlight differences in the performance of beat detection algorithms between different patient groups, which were observed not only for *MSPTDfast (v.2)* but also almost all other beat detectors. The *F*_1_-score of *MSPTDfast (v.2)* was significantly lower on AF subjects in comparison to non-AF subjects (see (a)); on neonates in comparison to adults (see (b)); and on White subjects compared to Black subjects (see (c)). Of these, only the difference in performance between adults and neonates remained significant after correcting for multiple comparisons.

**Figure 5.**
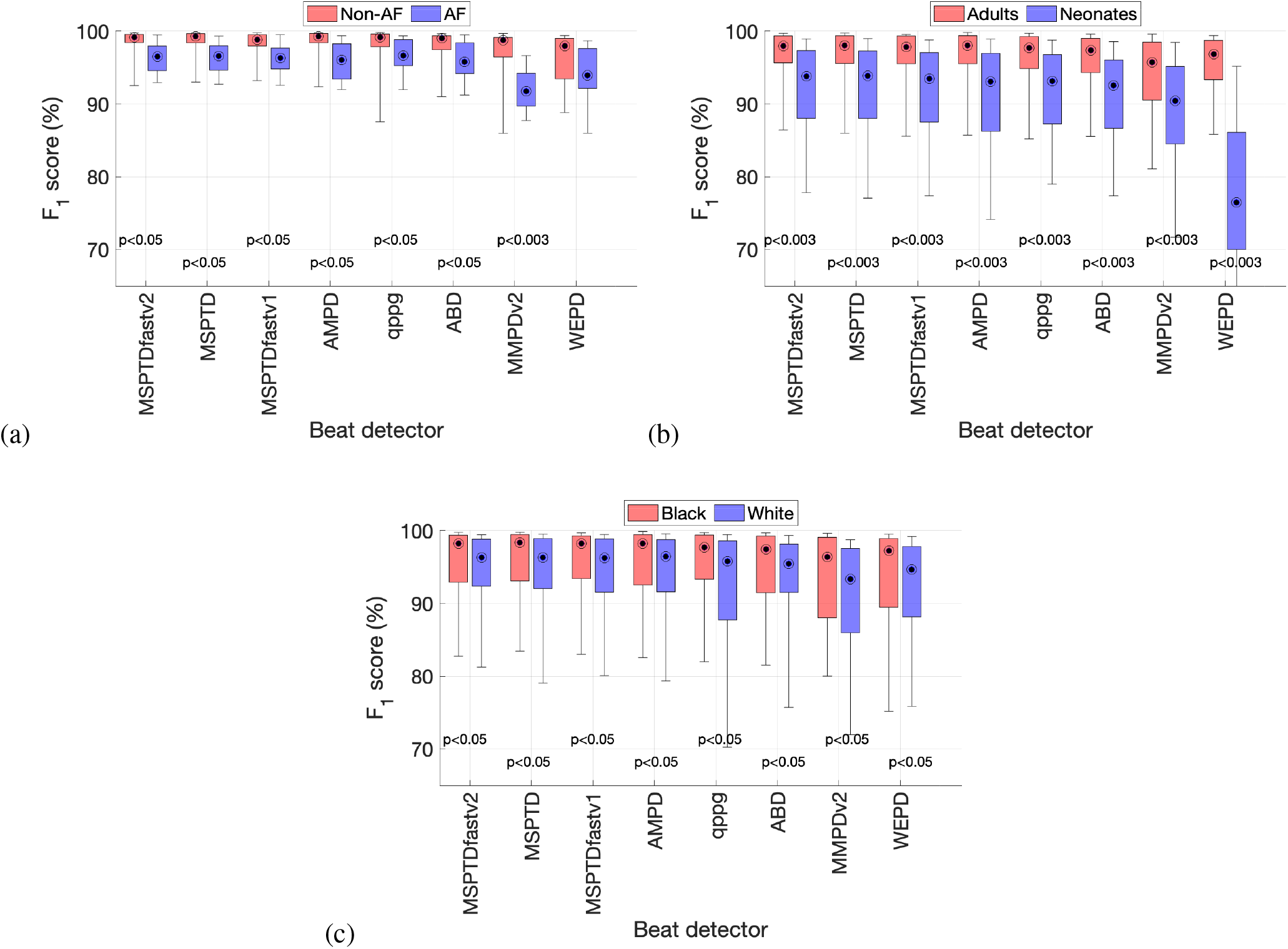
Associations between beat detection accuracy and patient characteristics: (a) atrial fibrillation (AF) vs. non-AF; (b) adults vs. neonates; and (c) Black vs. White subjects. p<0.05 indicates a significant difference before correction for multiple comparisons, whereas a p<0.003 indicates a significant difference after correction for multiple comparisons. Corresponding results for sensitivity and positive predictive value are shown in Appendix C.

## 4. Discussion

### 4.1. Summary of findings

This study reports the development and benchmarking of *MSPTDfast (v.2)*, an open-source PPG beat detection algorithm whose design is based on that of the *MSPTD* algorithm. The algorithm was found to have a much faster execution time than *MSPTD*, whilst maintaining a high *F*_1_-score. When compared to state-of-the-art beat detection algorithms on PPG data acquired in critical care and activities of daily living, *MSPTDfast (v.2)* was found to achieve the highest *F*_1_-score (alongside *MSPTD*) and one of the shortest execution times. This study indicates that *MSPTDfast (v.2)* is an accurate and efficient PPG beat detection algorithm. An open-source implementation is available in Matlab format.

The improved efficiency of *MSPTDfast (v.2)* in comparison to *MSPTD* was achieved by incorporating two key changes: (i) pre-processing PPG signals to reduce the sampling frequency to 20 Hz prior to analysis; and (ii) only calculating scalogram scales corresponding to heart rates >30 bpm. Both of these improvements reduce the computational complexity of calculating the scalogram matrices, which is the most computationally demanding part of the algorithm. Whilst efficiency could potentially have been improved further by making additional refinements (such as downsampling to 10 Hz instead of 20 Hz, using a shorter window duration of 4 s instead of 6 s, and detecting only either pulse peaks or onsets rather than both), we selected design options based on not only efficiency but also beat detection accuracy. Whilst there was no evidence that downsampling to 10 Hz or using a window duration of 4 s would have reduced accuracy in this study, we preferred the more conservative choices of 20 Hz and 6 s in an attempt to maintain accuracy in potential edge cases (namely very high or low heart rates).

The improvement in efficiency conferred by *MSPTDfast (v.2)* is closely related to the original sampling frequency of the PPG signal (see Figure 3(b)), as demonstrated by the greatest improvement in efficiency being observed on the CapnoBase dataset (in which PPG signals are sampled at 300 Hz), followed by the BIDMC and MIMIC PERform datasets (125 Hz), and finally PPG-DaLiA (64 Hz). Nonetheless, in all these cases the *MSPTDfast (v.2)* algorithm conferred a substantial improvement in efficiency compared to *MSPTD*.

This study confirmed key limitations of *MSPTDfast (v.2)* as well as other PPG beat detection algorithms. As observed in [6], most beat detection algorithms performed worse in neonates than adults. Not only was this the case for the original *MSPTD* algorithm, but it remained the case for the proposed *MSPTDfast (v.2)*. This is a key area for future work. In addition, poorer performance of *MSPTDfast (v.2)* was observed in AF signals compared to non-AF signals, although this difference did not remain significant after correction for multiple comparisons. Nonetheless, improving performance in AF may also indicate an area for future development [24].

### 4.2. Comparison with the literature

The development of *MSPTDfast (v.2)* builds on work spanning over a decade by many researchers, as illustrated in Figure 1. It benefited firstly from the original and subsequent work on algorithm design which laid the basis for the development of *MSPTDfast (v.2)* [8, 16–18]. *In addition, it benefited from a recent study benchmarking PPG beat detection algorithms [6]*, *which identified MSPTD* as a leading PPG beat detection algorithm and provided an open-source implementation. This development process was greatly aided by the provision of source code in some of the previous studies [6, 8, 16].

In this work a key step in improving the efficiency of *MSPTD* was to introduce a downsampling step, whereby the PPG signal was downsampled to approximately 20 Hz prior to analysis. This is a counter-intuitive step as devices often use much higher sampling frequencies than this to measure the PPG. Yet, it follows a trend in the literature which has observed that it is possible to estimate physiological parameters accurately from PPG signals at relatively low sampling frequencies, such as: (i) 5 Hz for heart rate estimation [34]; (ii) ≥ 16 Hz for respiratory rate estimation [35]; (iii) ≥ 25 Hz or 50 Hz for pulse rate variability analysis [36, 37]; and (iv) ≥ 60 Hz for pulse wave feature extraction [38]. Similarly, in this study, no reduction in performance was observed for beat detection when using signals of ≥ 20 Hz. Reducing the sampling frequency may reduce power consumption and therefore extend battery life by reducing the power used in sensing, data transmission and/or storage, and processing [39].

### 4.3. Strengths and limitations

The key strengths of this study are as follows. First, the proposed algorithm was developed and tested on several different datasets which are representative of different settings in which photoplethysmography is commonly used: both hospital monitoring and wearable monitoring in daily life. The use of several datasets provides confidence about the generalisability of the algorithm’s performance. Second, the proposed algorithm was benchmarked against several state-of-the-art algorithms, providing strong evidence for its high performance. Third, an open-source implementation of the algorithm was provided for ease of use.

The key limitations of the study are as follows. First, the different datasets used are not entirely independent. The eight datasets listed in Table 1 were derived from four original datasets (since five of the datasets were derived from the larger MIMIC database). In addition, the WESAD and PPG-DaLiA datasets were acquired using the same device from similar groups of subjects. Second, there can be a high level of heterogeneity between PPG signals, which was not fully captured by the datasets used in this study. For instance, signals can vary greatly between subjects (such as young and elderly subjects) and between devices (due to differences in factors such as body position, wavelength of light, and contact vs. non-contact imaging photoplethysmography). Third, in the benchmarking assessment we only included open-source beat detectors, and we do not know how the performance of the proposed algorithm would compare to proprietary algorithms (*e.g*. SWEPD, which was designed for accurate beat detection in AF [24]). Fourth, there are limitations to the use of execution time as a metric to assess algorithm efficiency, particularly when used on a computer rather than with target monitoring devices, as it may not capture differences in real-world efficiency [40] and reflects the execution time of an algorithm implementation rather than the intrinsic computational complexity of the algorithm itself [41]. Fifth, some of the comparator algorithms were implemented by ourselves rather than the original authors, so are subject to potential inaccuracies in our understanding or implementation of the algorithms. Finally, the proposed algorithm has not been designed for real-time use, and it may be possible to make further improvements for use on embedded devices, particularly given the large memory requirements of the scalograms (see [16] for further details).

### 4.4. Implications

The *MSPTDfast (v.2)* algorithm may be of great utility in research, particularly when conducting research with larger photoplethysmography datasets. We have tried to make the algorithm accessible to others by providing an open-source Matlab implementation. Furthermore, the algorithm is provided under the permissive MIT license to aid reuse (although as the license states - it is provided without warranty of any kind). Researchers may benefit not only from the open-source algorithm, but also from the open-science practices adopted in this study: (i) the datasets used are all freely available (although admittedly the WESAD and MIMIC PER-form Ethnicity dataset require some derivation using the provided open-source code); (ii) the PPG beat detection algorithms used are all open-source (*MSPTDfast (v.2)* and comparator algorithms); (iii) experiments were conducted using the open-source *ppg-beats* evaluation framework; and (iv) tutorials are provided to reproduce the work. These resources are all documented at https://ppg-beats.readthedocs.io/, and archived at https://zenodo.org/doi/10.5281/zenodo.6037646 [9].

Further research is required to determine the potential clinical utility of the *MSPTDfast (v.2)* algorithm. Key areas for future work include assessing the algorithm’s performance on other clinical datasets, and improving the algorithm’s performance in neonates. Furthermore, to improve clinical utility, the algorithm should be coupled with a quality assessment component that automatically assesses the confidence level of each beat detection. This would allow only beat detections with high confidence levels to be used in analyses that inform clinical decision making. This approach has been found to increase the accuracy of pulse oximetry-based heart rate monitoring [42].

More broadly, the growing body of evidence that physiological information can be extracted from PPG signals at relatively low sampling frequencies indicates that it may be beneficial to reduce the sampling frequency used by devices. This could be particularly beneficial in wearable devices, where it could increase battery life. PPG sensors have a particularly high power consumption in comparison to ECG or accelerometry sensors, making reducing their power consumption a key factor in the battery life of wearables when used for physiological monitoring. measurements. It may be possible to use sampling frequencies of 60 Hz or less to obtain physiological parameters accurately, depending on the parameters to be extracted. Indeed, some devices already use relatively low sampling frequencies, such as 25 Hz in the PulseOn Optical Heart Rate Monitor device [43], and 64 Hz in the Empatica E4 device [44]. The potential benefits of using more efficient algorithms aided by lower sampling frequencies are not only linked to the user acceptance of a device, but also its carbon footprint [45].

## 5. Conclusion

The *MSPTDfast (v.2)* algorithm presented in this study is an accurate and efficient PPG beat detection algorithm. It was found to be considerably more efficient than its predecessor, *MSPTD*, with no reduction in accuracy. *MSPTDfast (v.2)* is openly available in the Matlab *ppg-beats* tool-box.

## Data Availability

This study used publicly available data, which is available at:
- MIMIC PERform Datasets: https://zenodo.org/doi/10.5281/zenodo.6807402
- CapnoBase: https://doi.org/10.5683/SP2/NLB8IT
- BIDMC: https://doi.org/10.13026/C2208R
- WESAD: https://doi.org/10.24432/C57K5T
- PPG-DaLiA: https://doi.org/10.24432/C53890
The code used in the study is available at:
- https://doi.org/10.5281/zenodo.13368243
The accompanying documentation is available at:
- https://ppg-beats.readthedocs.io/en/latest/

## Acknowledgments

This study is funded by the British Heart Foundation [FS/20/20/34626]. ChatGPT (OpenAI, San Francisco, CA, USA) was used for language editing.

### Appendix

#### A. Additional internal validation results

Additional results for the sensitivity and positive predictive value of beat detection algorithms are shown in Figure 6.

**Figure 6.**
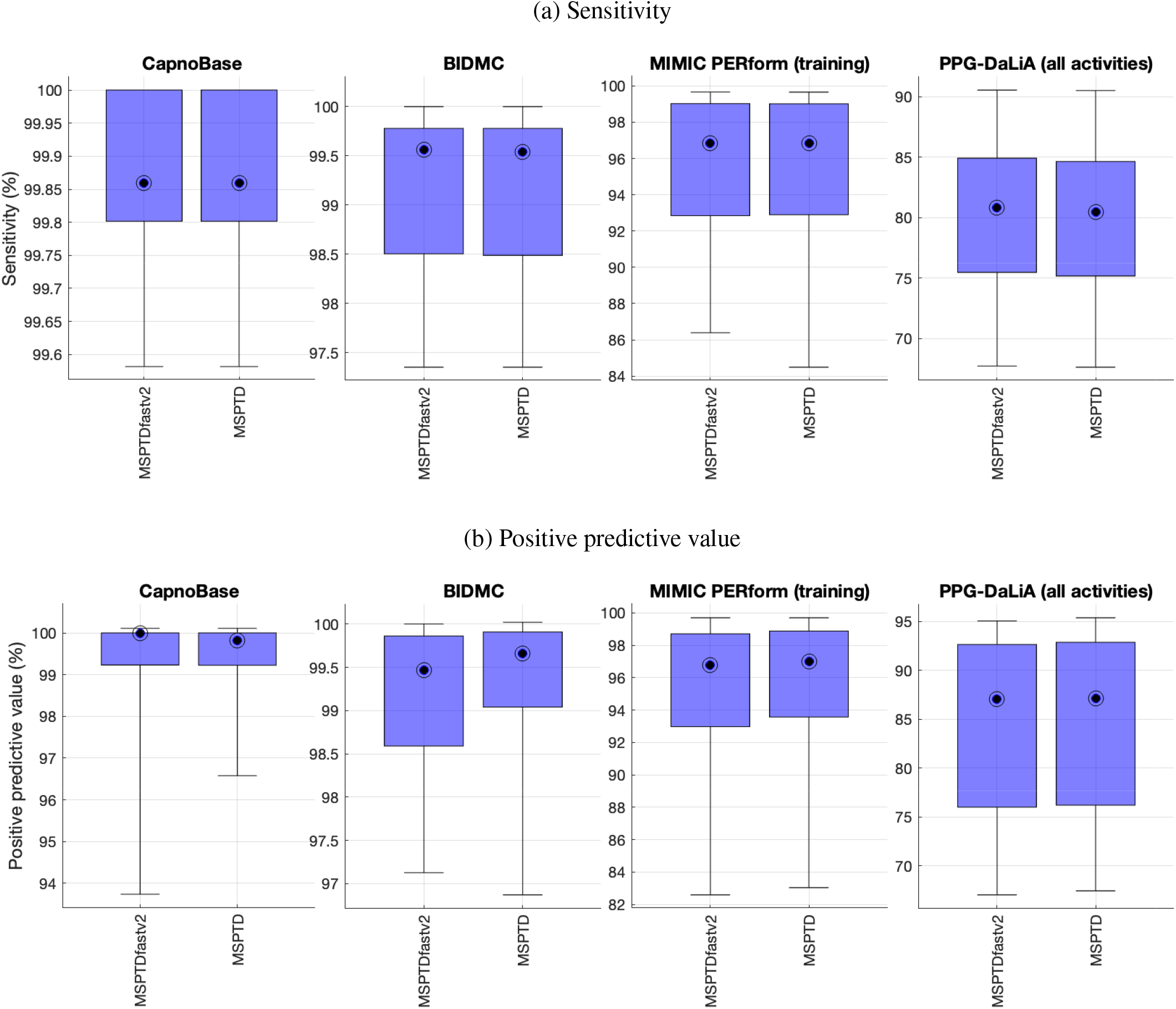
Internal validation of *MSPTDfast (v.2)* against *MSPTD* on the development datasets: additional results showing the sensitivity and positive predictive value of the algorithms

#### B. Additional benchmarking results

Additional results for the sensitivity and positive predictive value of beat detection algorithms are shown in Figure 7 and Table 5.

**Table 5.** Benchmarking *MSPTDfast (v.2)*against leading beat detection algorithms: additional results for the sensitivity and positive predictive value of the algorithms. Results are expressed as median (lower - upper quartiles). *Definition: * indicates a significant difference compared to MSPTDfast (v.2)*.

| Algorithm | MIMIC PERform (Testing) |  | WESAD (all activities) |  |
| --- | --- | --- | --- | --- |
|  | Sensitivity (%) | PPV (%) | Sensitivity (%) | PPV (%) |
| MSPTD | 97.4 (91.7-99.0) | 97.2 (92.0-99.0) * | 83.0 (80.2-86.1) * | 86.2 (82.1-90.4) * |
| MSPTDfastv2 | 97.2 (91.7-99.1) | 97.1 (91.7-98.9) | 83.0 (80.2-86.1) | 86.1 (81.9-90.2) |
| AMPD | 96.6 (90.5-98.8) * | 97.3 (91.7-99.1) * | 80.1 (77.7-83.4) * | 86.3 (82.5-90.1) * |
| MSPTDfastv1 | 97.1 (90.9-99.0) * | 97.0 (91.8-98.8) * | 82.5 (80.2-85.6) * | 86.2 (82.1-90.2) * |
| qppgfast | 96.1 (89.5-98.7) * | 96.8 (92.7-98.9) | 81.8 (77.2-85.3) * | 84.2 (78.5-88.8) * |
| ABD | 96.2 (88.7-98.6) * | 96.5 (91.6-98.7) | 81.0 (78.8-85.2) * | 84.6 (80.8-89.0) * |
| MMPDv2 | 92.5 (86.4-97.7) * | 95.9 (89.4-98.3) * | 85.5 (83.2-88.5) * | 83.7 (79.4-88.6) * |
| WEPD | 90.2 (65.2-97.9) * | 95.3 (90.4-98.2) * | 84.1 (81.5-86.4) * | 81.0 (78.0-88.5) * |

**Figure 7.**
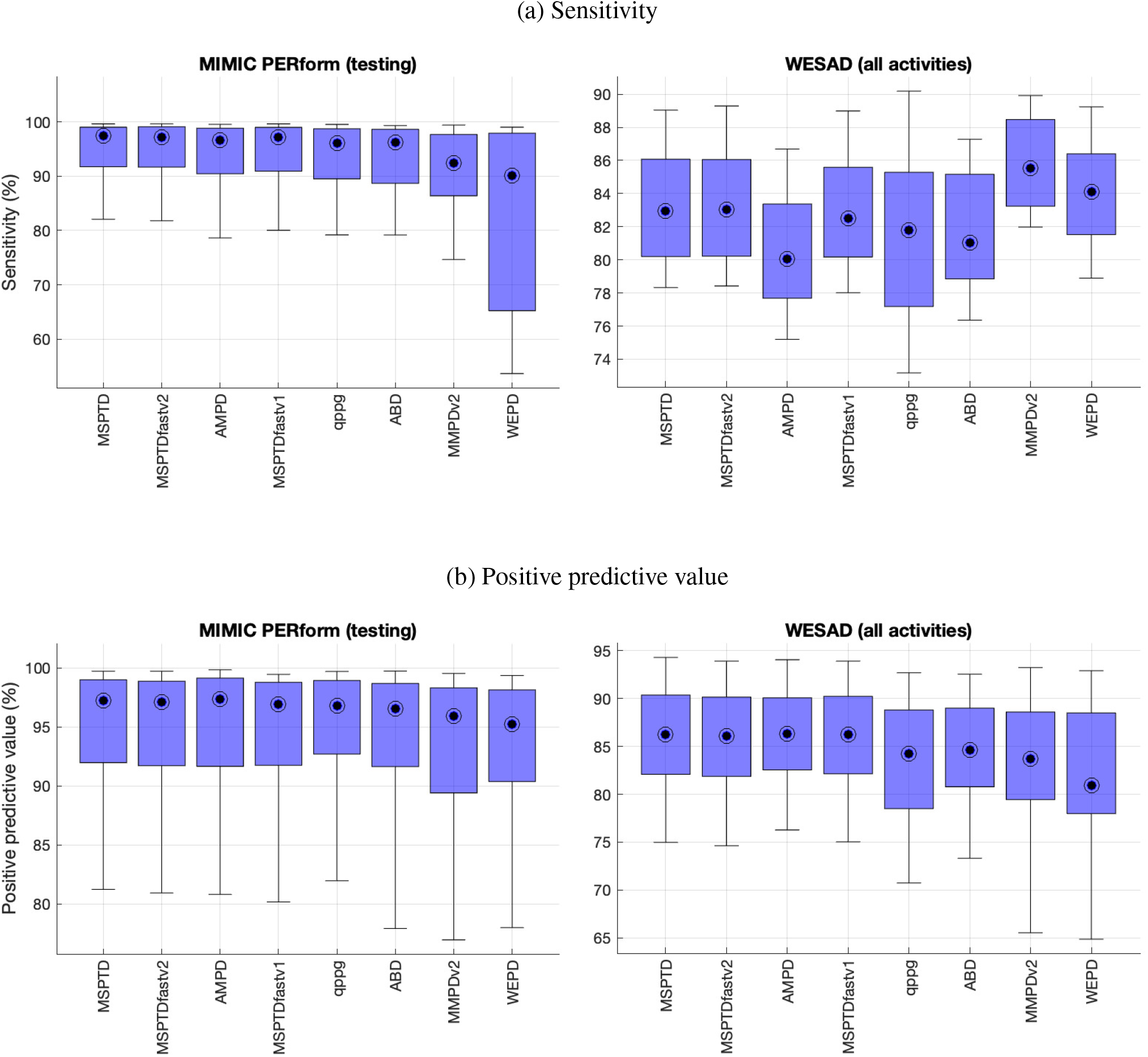
Benchmarking *MSPTDfast (v.2)* against leading beat detection algorithms: additional results showing the sensitivity and positive predictive value of the algorithms.

#### C. Additional results on associations between beat detection accuracy and patient characteristics

Additional results for the associations between beat detection accuracy and patient characteristics in terms of sensitivity and positive predictive value are shown in Figure 8.

**Figure 8.**
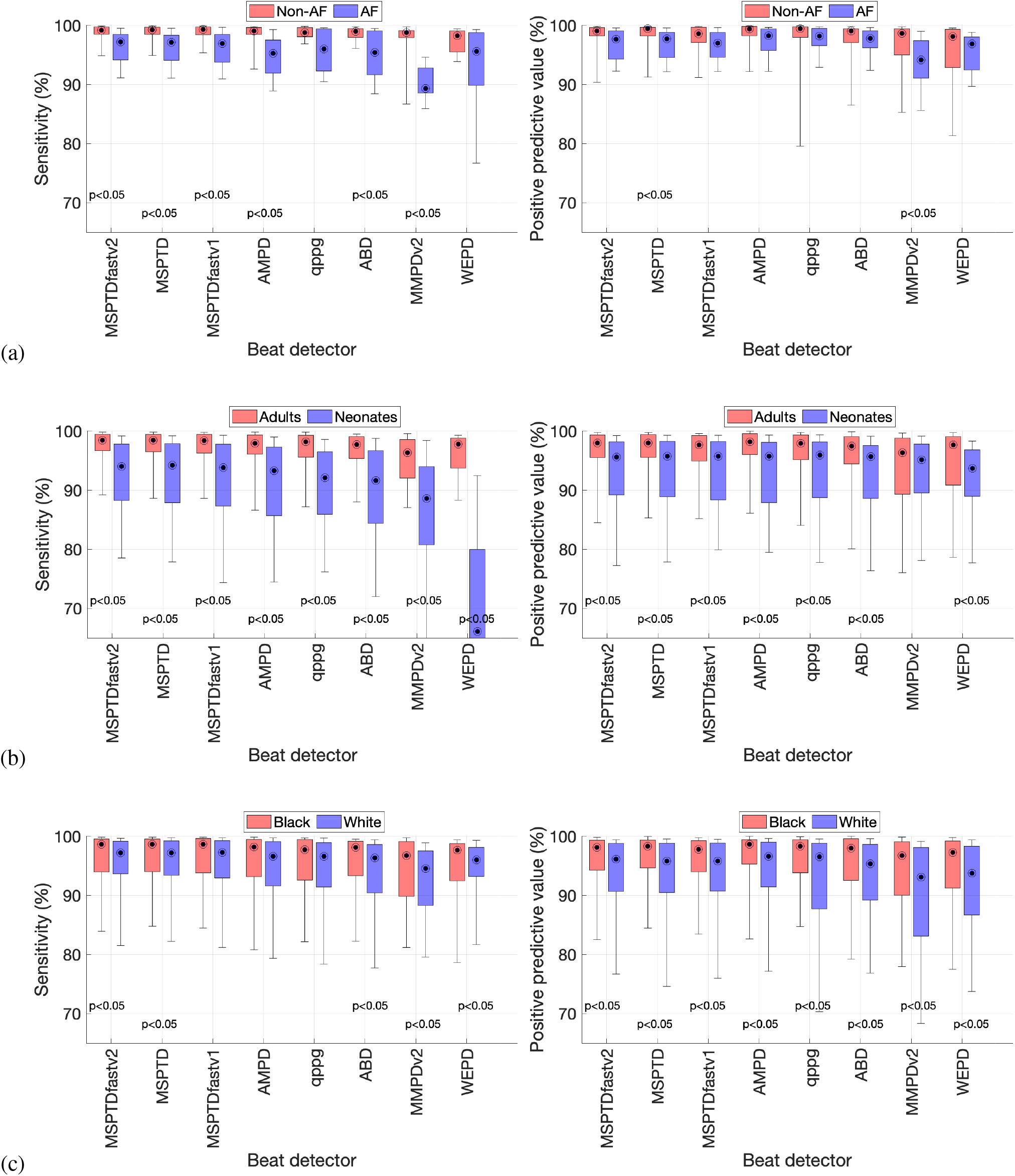
Associations between beat detection accuracy and patient characteristics: (a) atrial fibrillation (AF) vs. non-AF; (b) adults vs. neonates; and (c) Black vs. White subjects. p<0.05 indicates a significant difference (no correction was made for multiple comparisons in this analysis).

## Footnotes

1 In [6] this algorithm was named the ‘Peak Detection Algorithm’, PDA.

